# Digital inclusion, access barriers and trust calibration in smartphone-based hypertension screening: a mixed-methods policy and implementation study in northern Nigeria

**DOI:** 10.64898/2026.08.07.26359947

**Authors:** David Dasa, Philip Davies

## Abstract

**Objectives:** To assess how digital inclusion factors and physical access barriers are associated with user trust in smartphone-based remote photoplethysmography (rPPG) hypertension screening, and to identify implications for digital health policy, procurement and implementation in low-resource settings.

**Methods:** Cross-sectional mixed-methods survey in five outpatient clinics in Kebbi State, northern Nigeria (*N* =287). Trust was measured using comfort, confidence and perceived usefulness Likert scales. Primary analyses used binary logistic models with HC3 robust standard errors; sensitivity analyses are reported in supplementary material. Free-text responses were thematically analysed.

**Results:** Smartphone ownership was 51.2%; Transsion-brand devices comprised 56.5% of owners. Greater distance to a blood pressure facility was independently associated with lower perceived usefulness (OR 0.51, 95% CI 0.30–0.87; *p*=0.013) and lower comfort (OR 0.61, 0.37–0.98; *p*=0.042). Among owners, Transsion versus Samsung showed higher confidence odds (OR 3.82, 1.02–14.27; *p*=0.046). Qualitative themes supported the implementation interpretation: platform-fit and device speed requests among Transsion owners; connectivity and offline-first concerns among those with greater travel distance. No brand contrast achieved FDR-adjusted significance; brand findings are exploratory.

**Conclusions:** Digital health policy and health technology assessment for smartphone-based screening should incorporate local device ecology, connectivity constraints, physical access burden and trust-calibration safeguards. Pre-implementation assessment of these factors is necessary for equitable and safe rPPG adoption in low-resource health systems.

**Public Interest Summary:** Smartphone-based blood pressure screening could improve access to hypertension services in low-resource settings, but safe implementation depends on more than technical accuracy. In five outpatient clinics in northern Nigeria, we found that user trust in remote photoplethysmography was shaped by smartphone access, device brand familiarity and distance from existing blood pressure services. Transsion Android devices—Tecno, Infinix and Itel—were the dominant smartphone type among owners, while users further from blood pressure facilities raised more concerns about internet access and offline use. These findings suggest that digital health policies should not assume a single smartphone-based screening tool will work equally well for all populations. Before implementation, health systems should assess local device ecology, connectivity, access barriers and the need for cuff-based confirmation, so that enthusiasm for new tools does not outpace safe clinical use.

## 1 Introduction

Hypertension accounts for a disproportionate burden of cardiovascular mortality in sub-Saharan Africa, yet the infrastructure for routine screening is constrained by workforce limitations, facility access barriers and travel burden in many low-resource settings [2]. Remote photoplethysmography (rPPG)—a face-capture technology using smartphone cameras to estimate blood pressure proxies—is being evaluated as a low-cost approach to extending screening reach in resource-constrained settings [1, 3]. Digital screening technologies offer genuine promise for expanding coverage, but adoption decisions based primarily on technical promise, without accounting for local implementation realities, risk widening health inequities or generating unsafe clinical behaviour.

A critical implementation risk is the perception–performance gap: users frequently rate digital health tools favourably despite moderate or weak validated accuracy [4, 5]. In AI-assisted clinical tools more broadly, this overconfidence has been shown to accelerate adoption ahead of evidence [5]. For rPPG specifically, trust calibrated to brand familiarity or access convenience rather than clinical performance constitutes a patient safety and governance risk, particularly where clinicians and patients may substitute rPPG readings for cuff-based measurement before validation thresholds are met. Digital health policy frameworks must therefore address not only technical accuracy but also the structural drivers of user trust.

Two structural driver classes are especially salient in low-resource settings. First, digital inclusion—device ownership and brand familiarity—may function as a trust heuristic in digitally stratified populations [6, 8, 9]. In many LMIC markets, Transsion-brand devices (Tecno, Infinix, Itel) dominate smartphone sales. In our study setting in northern Nigeria, Transsion devices represent 56.5% of smartphone ownership while Samsung accounts for only 9.5%—a device ecology with direct implications for procurement and validation strategy: rPPG tools tested on premium handsets may perform differently on locally dominant hardware. Second, physical access barriers—operationalised as distance to an existing blood pressure facility—may alter perceived usefulness and trust calibration by interacting with connectivity, power reliability and workflow constraints [10, 11]. Users facing high travel burdens may value decentralised access, but they may also be more sceptical if remote tools do not solve the practical barriers they experience.

This paper contributes policy-relevant evidence on three dimensions: (1) the device ecology surrounding smartphone-based screening in a representative LMIC clinical setting; (2) how physical access burden is associated with perceived usefulness; and (3) trust-calibration signals that should inform implementation safeguards. We conducted a cross-sectional mixed-methods survey in five outpatient clinics to quantify these associations and to triangulate quantitative findings with qualitative implementation themes. A conceptual framework linking these dimensions to policy levers is presented in Box 1.

### Box 1. Conceptual framework for safe implementation of smartphone-based rPPG screening in low-resource settings.

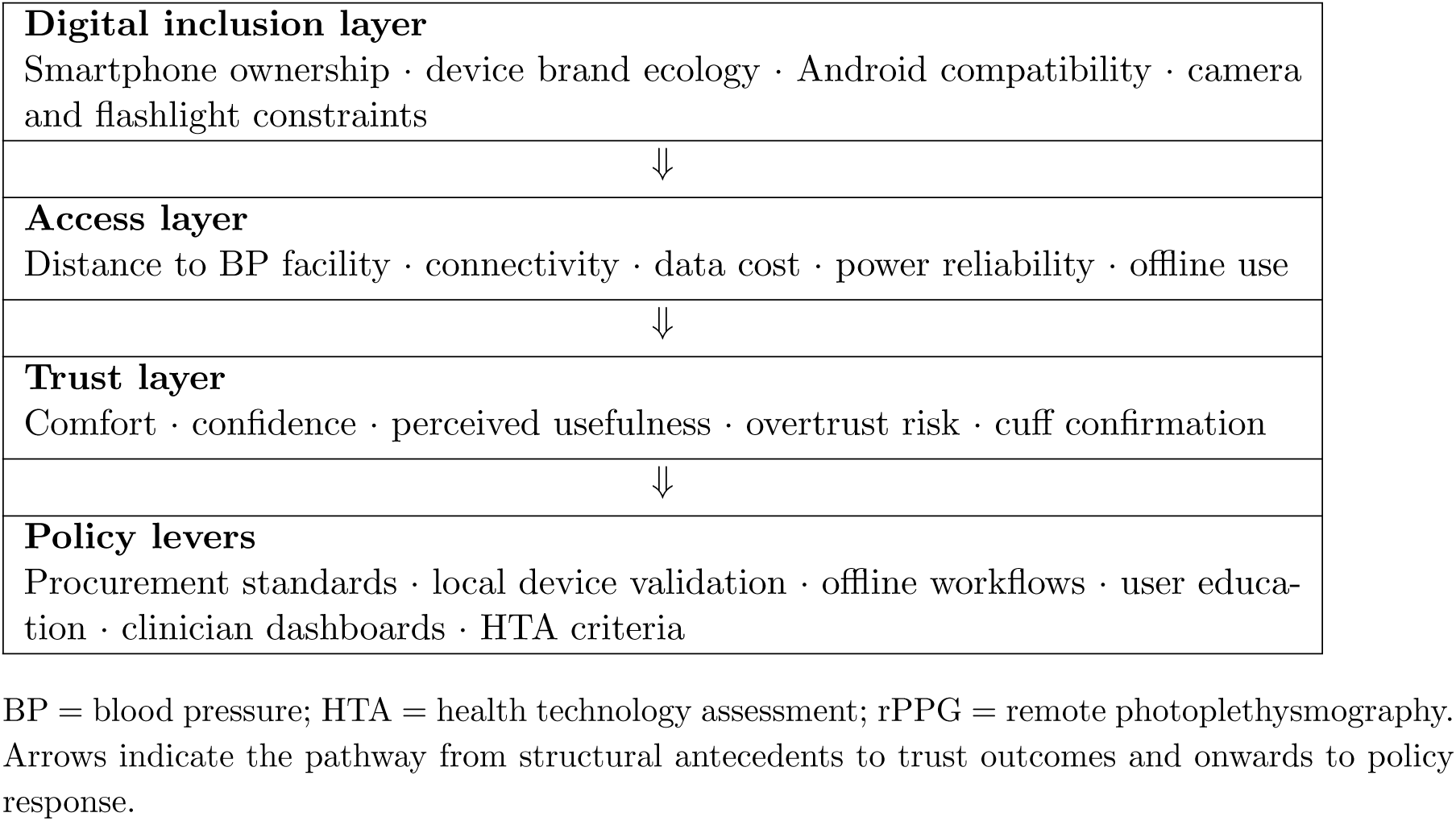

## 2 Methods

### 2.1 Design, setting and research questions

We conducted a cross-sectional mixed-methods survey in five outpatient clinics in Kebbi State, northwestern Nigeria, following STROBE reporting guidelines [12]. The study was embedded within a larger multi-site rPPG field evaluation [4]. Eligible participants were adult patients and clinical staff who used a face-capture rPPG application during routine outpatient attendance.

The study addressed three policy-facing research questions: (1) How are digital inclusion factors associated with user trust in smartphone-based hypertension screening? (2) How does physical access to existing blood pressure services relate to perceived usefulness? (3) What implementation signals emerge that should inform procurement, workflow design and governance?

### 2.2 Measures

#### Outcomes

Comfort, confidence, and perceived usefulness were each measured on a five-point Likert scale (1 = strongly disagree, 5 = strongly agree). Responses were dichotomised as *top-box* (5/5 versus all else), representing unambiguous endorsement consistent with adoption-threshold language used in health technology assessment.

#### Smartphone ownership and brand

Participants reported smartphone ownership (Yes/No). Among owners, brand was classified from structured entries and free-text: Transsion (Tecno, Infinix, Itel combined), Samsung, iPhone, Other-Android, or Unknown. Itel was identified via free-text parsing and assigned to Transsion based on corporate ownership.

#### Distance to a blood pressure facility

Participants answered: “How far do you live from a place you can check blood pressure?” with ordered options coded 0–3: **0** within my compound/home; **1** within my street or very nearby; **2** within my town but not nearby; **3** outside my town or requires travel. Missing values were excluded from distance models (full sample *n*=9; owners *n*=8).

### 2.3 Statistical analysis

Binary logistic models for top-box (5/5) outcomes with HC3 heteroskedasticity-robust standard errors were the primary analyses [16]. Brand-group effects were estimated among owners only (Samsung as reference); ownership effects were estimated in the full sample without brand terms. Covariates were age (years) and gender; an ownership × distance interaction was included in the ownership model. Cumulative logit models for five-level outcomes [13] and Firth bias-reduced logistic models for sparse brand cells [15] were conducted as sensitivity analyses and are reported in supplementary material, along with Benjamini–Hochberg FDR *q*-values [14]. Main-text *p*-values are descriptive.

### 2.4 Qualitative analysis

Free-text responses were analysed thematically [22]. Two coders independently piloted 20% of entries, refined the codebook iteratively, then double-coded a random 10% subsample blinded to quantitative results [19]. Three prespecified domains guided coding: *Digital Inclusion* (Android availability, device speed, brand fit), *Access Constraints* (travel burden, connectivity, offline needs, power reliability), and *Trust Calibration* (accuracy concerns, cuff confirmation, privacy). Inter-rater reliability was computed via observed agreement (*P_o_*), Cohen’s *κ*, Gwet’s AC1, and PABAK [20, 21].

### 2.5 Ethics

Ethical approval was obtained from the Kebbi State Health Research Ethics Committee (107:017/2024) and Bournemouth University Research Ethics Committee (ID 55993), in accordance with the Declaration of Helsinki. Written informed consent was obtained from all participants. Participants with incidentally detected hypertension were referred according to local care pathways.

## 3 Results

### 3.1 Implementation context: smartphone ownership, device ecology and access burden

A total of 287 participants were enrolled. Smartphone ownership was 51.2% (*n*=147), indicating that approximately half the target population would be structurally excluded from any exclusively smartphone-based screening pathway at point of deployment. Among owners, the Transsion group (Tecno 35.4%, Infinix 19.0%, Itel 2.0%) comprised 56.5% of the device ecology; Samsung owners represented 9.5% (Table 1, Figure 1). This Transsion-dominant, premium-brand-marginal ecology reflects the broader LMIC smartphone market in the region and constitutes a direct procurement and validation consideration for any smartphone-based screening programme.

**Figure 1:**
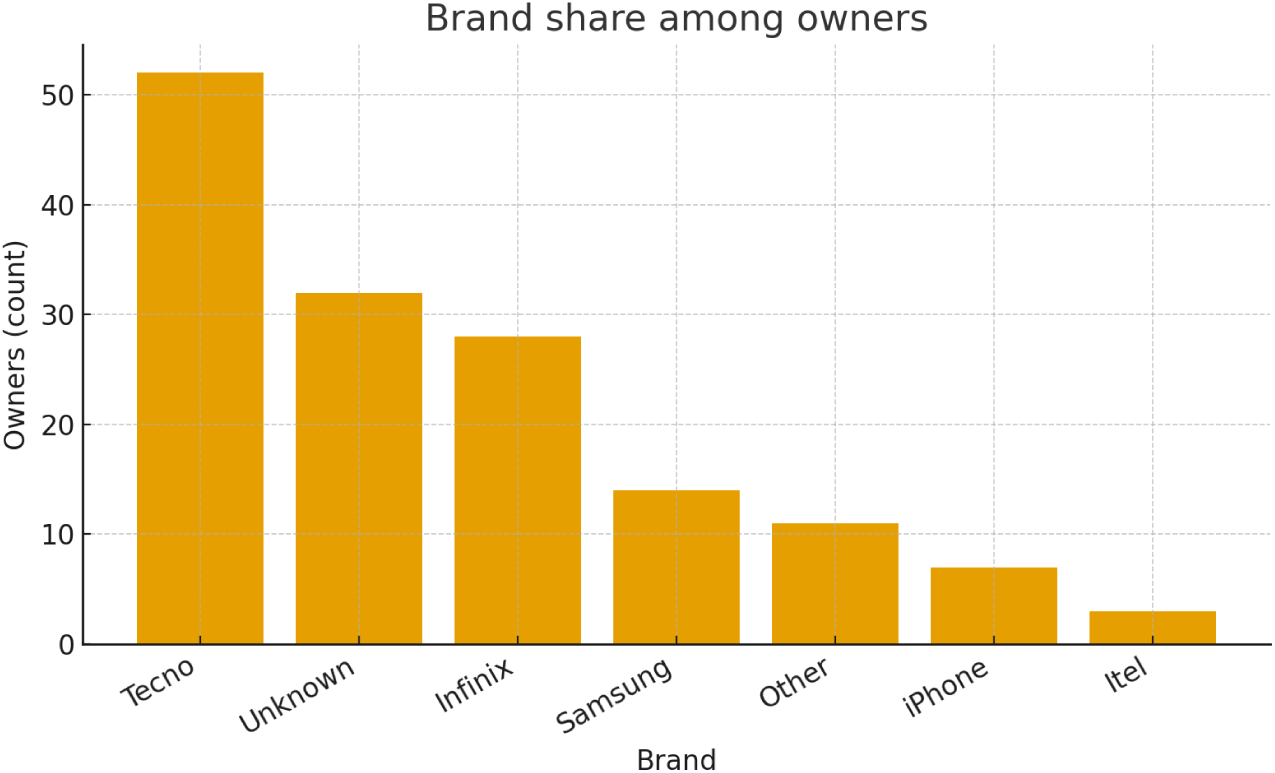
Brand share among smartphone owners. Transsion (Tecno, Infinix, Itel) constitutes the majority of the owner sample, reflecting the dominant device ecology in the study region. This distribution is a procurement and validation consideration for smartphone-based screening programmes.

**Table 1:** Smartphone ownership and brand distribution among owners (*n*=147).

| Brand group | Count | % of owners |
| --- | --- | --- |
| Transsion total (Tecno + Infinix + Itel) | 83 | 56.5 |
| Tecno | 52 | 35.4 |
| Infinix | 28 | 19.0 |
| Itel | 3 | 2.0 |
| Samsung | 14 | 9.5 |
| iPhone | 7 | 4.8 |
| Other-Android | 11 | 7.5 |
| Unknown | 32 | 21.8 |

Distance to a BP facility: within compound (*n*=3), nearby street (*n*=68), within town (*n*=100), outside town (*n*=107), missing (*n*=9). Among valid responses (*n*=278), 74.5% lived within town but not nearby or outside town; 38.5% reported that BP measurement was outside their town or required travel—the highest distance category on the four-level scale.

### 3.2 Adoption-relevant perceptions by ownership and brand group

Top-box adoption-relevant endorsement was consistently higher among owners than non-owners (Figure 2): perceived usefulness 66.4% vs 45.5%; comfort 56.2% vs 45.0%; confidence 49.7% vs 31.8%. Among owners, Transsion and iPhone/Other-Android groups showed higher top-box usefulness than Samsung, though iPhone and Other-Android cells were sparse (Figure 2b).

**Figure 2:**
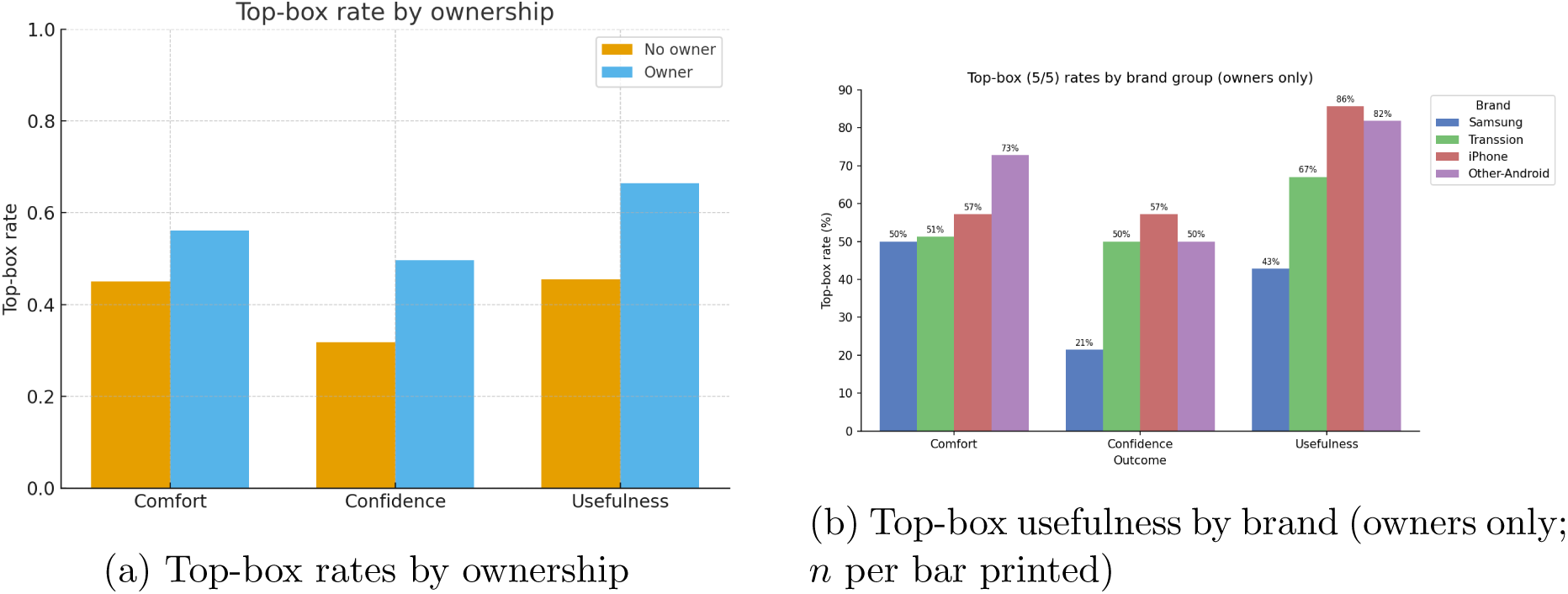
Unadjusted top-box (5/5) adoption-relevant endorsement rates by ownership and by brand among owners. Bars print group size to make cell sparsity explicit.

### 3.3 Adjusted associations with comfort, confidence and perceived usefulness

Adjusted odds ratios are given in Table 2. In the full-sample ownership model, distance was inversely associated with top-box usefulness (OR 0.51, 95% CI 0.30–0.87; *p*=0.013) and comfort (OR 0.61, 0.37–0.98; *p*=0.042). Age was negatively associated with all three top-box outcomes (*p*≤0.010). The smartphone ownership effect was not significant after adjustment, and the ownership × distance interaction was not significant across outcomes.

**Table 2:** Adjusted odds ratios (OR) for top-box (5/5) outcomes. HC3 robust 95% CIs and *p*-values throughout. Bold denotes *p<*0.05.

| Predictor | Comfort OR [95% CI]; $p$ | Confidence OR [95% CI]; $p$ | Usefulness OR [95% CI]; $p$ |
| --- | --- | --- | --- |
| *Ownership model (full sample; $n$: Comfort 248, Confidence 246, Usefulness 247)* |  |  |  |
| Smartphone ownership (Yes vs No) | 0.64 [0.22, 1.82]; 0.398 | 1.54 [0.54, 4.38]; 0.423 | 0.98 [0.33, 2.89]; 0.969 |
| Distance to BP facility (per step) | **0.61 [0.37, 0.98]; 0.042** | 1.11 [0.65, 1.88]; 0.709 | **0.51 [0.30, 0.87]; 0.013** |
| Ownership × Distance | 1.30 [0.68, 2.50]; 0.434 | 0.89 [0.45, 1.78]; 0.750 | 1.20 [0.59, 2.43]; 0.614 |
| Age (years) | **0.97 [0.95, 0.99]; 0.001** | **0.97 [0.95, 0.99]; 0.003** | **0.98 [0.96, 0.99]; 0.010** |
| Gender: Male vs Female | 1.22 [0.69, 2.15]; 0.500 | 1.35 [0.77, 2.37]; 0.302 | 1.39 [0.78, 2.48]; 0.258 |
| *Brand model (owners only; Samsung reference; $n$: Comfort/Usefulness 106, Confidence 105)* |  |  |  |
| Brand: Transsion vs Samsung | 1.27 [0.37, 4.29]; 0.704 | **3.82 [1.02, 14.27]; 0.046** | 3.36 [0.88, 12.79]; 0.076 |
| Brand: iPhone vs Samsung | 1.27 [0.19, 8.27]; 0.803 | 5.14 [0.67, 39.64]; 0.116 | **9.58 [1.19, 76.82]; 0.033** |
| Brand: Other-Android vs Samsung | 4.43 [0.74, 26.40]; 0.102 | 4.30 [0.70, 26.59]; 0.116 | **19.74 [1.44, 269.87]; 0.025** |
| Distance to BP facility (per step) | 0.75 [0.43, 1.29]; 0.297 | 1.13 [0.67, 1.93]; 0.639 | **0.53 [0.28, 0.99]; 0.046** |
| Age (years) | 0.99 [0.95, 1.03]; 0.582 | 0.99 [0.96, 1.03]; 0.684 | 1.02 [0.98, 1.06]; 0.335 |
| Gender: Male vs Female | 1.75 [0.76, 4.00]; 0.188 | 1.54 [0.68, 3.47]; 0.297 | 1.22 [0.51, 2.95]; 0.653 |

Among owners, Transsion versus Samsung showed higher confidence odds (OR 3.82, 95% CI 1.02–14.27; *p*=0.046) and a positive trend for usefulness (OR 3.36, 0.88–12.79; *p*=0.076). Distance retained its inverse association with usefulness among owners (OR 0.53, 0.28–0.99; *p*=0.046). Because brand-specific cells were small and no brand contrast survived FDR adjustment (Supplementary Table S2), brand findings should be interpreted as exploratory implementation signals rather than confirmatory evidence of brand-specific effects. Firth bias-reduced sensitivity analyses yielded estimates of the same sign and broadly similar magnitude.

Figure 3 shows predicted top-box usefulness probabilities across distance levels by ownership and by brand; the Transsion line tracks above Samsung at all distances.

**Figure 3:**
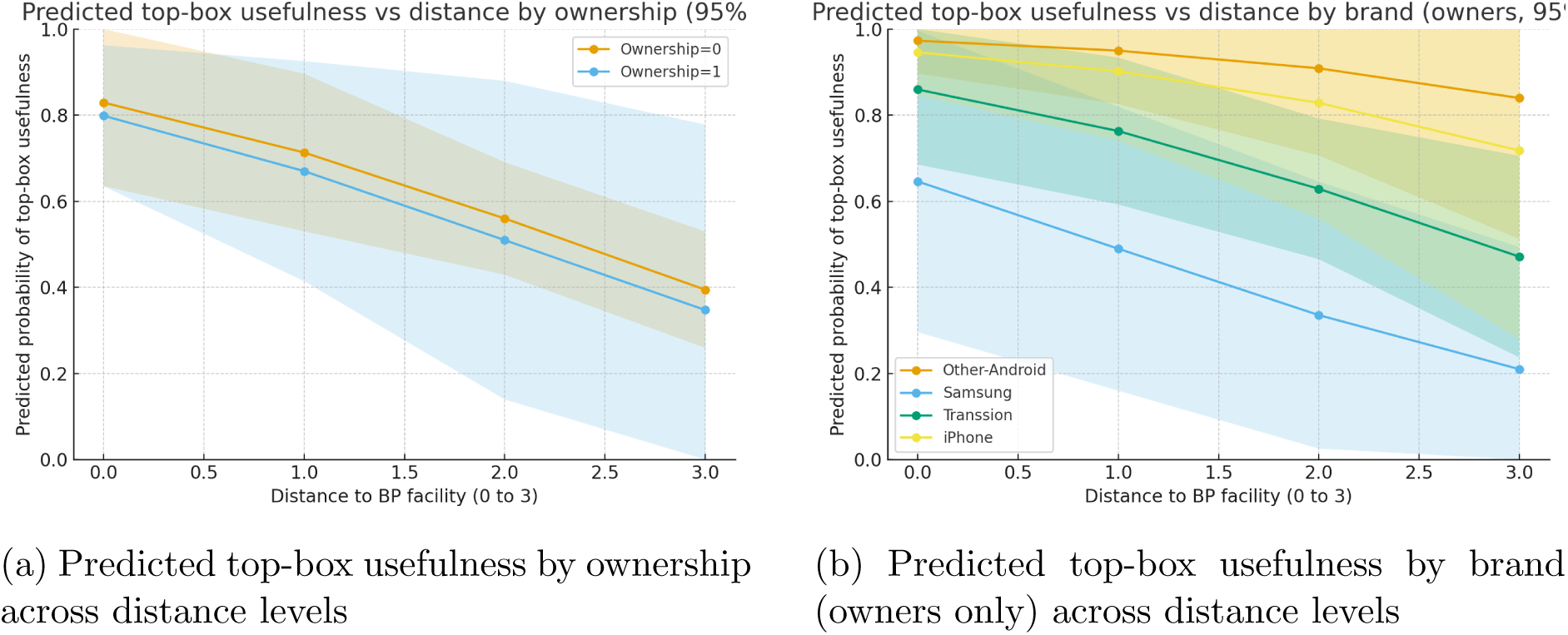
Predicted probability of top-box usefulness with 95% CI ribbons from primary logistic models. Shorter travel distance is consistently associated with higher predicted probability. Among owners, the Transsion line tracks above Samsung across all distance levels.

### 3.4 Qualitative evidence on implementation barriers and trust calibration

Inter-rater reliability on the double-coded subsample (*n*=26) was good-to-excellent across all three domains: *P_o_* 0.73–0.96; *κ* 0.46–0.91; AC1 0.47–0.94; PABAK 0.46–0.92.

Theme frequency counts by ownership and brand are in Table 3 and Figure 4. Owners generated substantially more digital inclusion content than non-owners (Android availability, device speed, capture affordances). Among owners, Transsion users concentrated requests on Android build quality, processing speed, flashlight/night-mode capture and hand placement guidance—themes consistent with platform fit as an explanation for their higher confidence odds and directly relevant to implementation design for Transsion devices. Samsung owners more frequently raised accuracy concerns. Connectivity and offline-first needs clustered among non-owners and those with greater travel distance, indicating that infrastructure constraints are a recognised barrier among the users most dependent on alternative access pathways.

**Figure 4:**
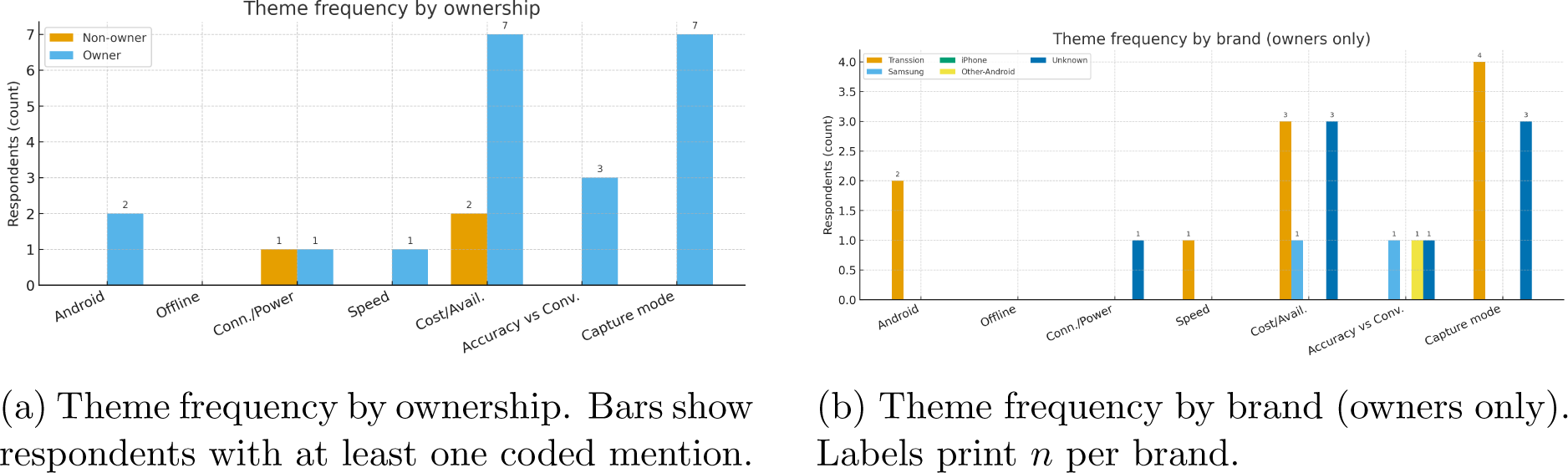
Qualitative implementation theme frequency by ownership and by brand. Digital inclusion content concentrates among owners and Transsion users; connectivity and offline needs concentrate among non-owners and those with greater travel distance.

**Table 3:** Qualitative theme counts by ownership and by brand (owners only). Counts are respondents with at least one coded mention.

| Group | Android | Offline | Conn./Power | Speed | Cost/Avail. | Accuracy vs Conv. | Capture mode |
| --- | --- | --- | --- | --- | --- | --- | --- |
| Non-owner | 0 | 0 | 1 | 0 | 2 | 0 | 0 |
| Owner | 2 | 0 | 1 | 1 | 7 | 3 | 7 |

Panel B. By brand (owners only)
| Brand group | Android | Offline | Conn./Power | Speed | Cost/Avail. | Accuracy vs Conv. | Capture mode |
| --- | --- | --- | --- | --- | --- | --- | --- |
| Transsion | 2 | 0 | 0 | 1 | 3 | 0 | 4 |
| Samsung | 0 | 0 | 0 | 0 | 1 | 1 | 0 |
| iPhone | 0 | 0 | 0 | 0 | 0 | 0 | 0 |
| Other-Android | 0 | 0 | 0 | 0 | 0 | 1 | 0 |
| Unknown | 0 | 0 | 1 | 0 | 3 | 1 | 3 |

Themes: Android availability; Offline-first needs; Connectivity/data/power; Device speed; Cost/availability; Accuracy vs convenience trade-off; Capture mode and affordances.

### 3.5 Mixed-methods integration for policy and implementation

Table 4 presents the quant–qual joint display for the three key predictors. Table 5 maps each predictor to its qualitative corroboration, the policy inference drawn, and the corresponding implementation implication for health systems and HTA. The convergence of quantitative estimates and qualitative themes across all three predictors strengthens the policy interpretation: device ecology, access burden and structural non-ownership are not statistical artefacts but embedded implementation challenges requiring explicit policy responses.

**Table 4:**
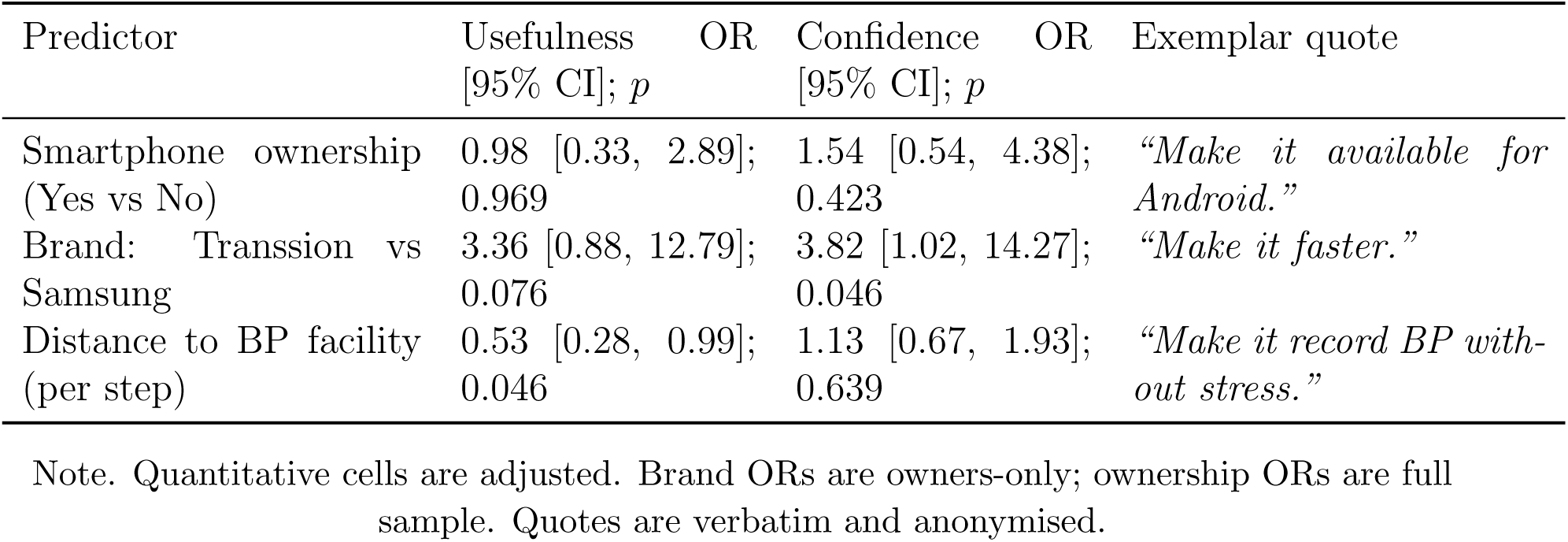
Quant–qual joint display for key predictors.

**Table 5:** Mixed-methods integration matrix: quantitative predictors, qualitative evidence, policy inference and implementation implication.

| Predictor (quant finding) | Qualitative theme | Policy inference | Implementation implication |
| --- | --- | --- | --- |
| Transsion > Samsung for confidence (OR 3.82; exploratory) | Platform fit, speed requests, capture affordances (Transsion owners) | Brand familiarity and Android fit act as trust heuristics; device ecology shapes adoption independently of accuracy | Validate rPPG builds on Transsion hardware before deployment; require cuff confirmation at first use; do not assume premium device performance extends to dominant local brands |
| Greater distance lowers usefulness (OR 0.51 per step; $p=0.013$) | Offline-first and connectivity concerns; convenience-oriented remarks | Access barriers recalibrate the performance users care about; participants with greater physical access burden may require stronger evidence of practical utility, offline functionality and confirmatory pathways before perceiving rPPG as useful | Offline-capable workflows; power-reliable deployment; targeted messaging for high-access-burden users; differentiated HTA assessment by access stratum |
| Ownership 51.2%; own-ership effect attenuates after adjustment | Owners raise usability and capture-aid requests; non-owners raise connectivity and cost barriers | Structural non-ownership constraints equitable reach from deployment day one | Hybrid care pathways; facility-based device access for non-owners; ownership and equity monitoring built into implementation frameworks |

Illustrative verbatim excerpts: *“make use of device flash light”* (patient, female, 25–34, Transsion owner); *“improve accuracy above current standard BP device”* (patient, male, 15–24, Samsung owner); *“make it work without need for internet”* (patient, male, 15–24, brand unknown).

## 4 Discussion

### 4.1 Principal findings

This study demonstrates that user trust in rPPG screening is structured by digital inclusion and physical access in ways that operate independently of the technology’s validated accuracy. The Transsion–Samsung confidence differential (OR 3.82) suggests brand familiarity functions as a trust heuristic: Transsion’s dominant market position means its rPPG implementation may feel more contextually credible. The inverse distance–usefulness relationship (OR 0.51 per step) indicates that physical access burden may reduce confidence in the practical utility of rPPG, likely because travel burden coexists with connectivity, power and workflow constraints. Rather than assuming that participants with greater physical access burden will automatically value remote screening more, implementation should treat this group as requiring offline workflows, clear triage messaging and confirmatory cuff access. These structural drivers, and the overtrust risk they generate, are the core policy findings of this study.

### 4.2 Implications for digital health policy

These findings carry direct implications for digital health policy in low-resource settings. The Transsion-dominated device ecology is not merely a technical variable but a procurement and validation criterion: programmes that recommend or procure rPPG tools validated only on premium handsets will reach users through devices for which clinical performance is unverified. Digital health policy frameworks should therefore mandate pre-implementation assessment of local device ecology—including dominant brands, Android version distribution and camera sensor specifications—as a standard condition for procurement approval or programme endorsement.

The 51.2% smartphone ownership rate also means that any exclusively smartphone-based screening pathway would structurally exclude approximately half the target population from deployment day one. Policy responses might include health-worker-mediated screening using facility devices, community access points, or hybrid cuff/smartphone pathways with clear referral protocols for non-owners. The policy implication is not that Transsion devices inherently cause higher trust, but that local device ecology must be measured before deployment and incorporated into validation, procurement and onboarding to prevent brand familiarity from substituting for evidence-based endorsement.

The WHO Global Strategy on Digital Health identifies equity, access and governance as core pillars of safe digital health deployment [23]. Our findings operationalise what those pillars require in practice for smartphone-based rPPG: a pre-deployment audit of device ownership, brand ecology and connectivity built into programme design rather than retrofitted after scale-up.

### 4.3 Implications for HTA and procurement

Standard health technology assessment frameworks evaluate diagnostic accuracy, cost-effectiveness and clinical impact. For smartphone-based rPPG in LMIC settings, this framework requires extension. A complete HTA should integrate: (a) technical accuracy validated across locally dominant device types; (b) equity of access by ownership and connectivity status; (c) offline workflow feasibility and cost; (d) implementation, training and compatibility testing costs; and (e) monitoring mechanisms for overtrust and inappropriate cuff substitution. These criteria do not require a full cost-effectiveness model at early deployment stage, but they do require that procurement decisions be evidence-informed rather than driven solely by headline diagnostic accuracy figures.

The inverse association between distance and perceived usefulness (OR 0.51 per step) presents a nuanced HTA challenge: users most in need of decentralised screening—those with the greatest travel burden—also express the most calibrated scepticism about rPPG utility compared with more proximate users. This may reflect realistic access awareness rather than disengagement, and suggests that implementation targeting and messaging strategies should be differentiated by access burden rather than uniform across populations.

### 4.4 Trust calibration as digital health governance

The trust–performance gap identified here—where enthusiasm outpaces validated accuracy [4, 5]—constitutes a governance challenge with patient safety implications. Trust calibrated to device familiarity or access convenience rather than clinical performance increases the risk that users substitute rPPG readings for validated cuff-based measurement before accuracy thresholds are met. Implementation governance should embed trust-calibration mechanisms as standard: mandatory cuff-based confirmation at first use and at regular intervals; clinician-facing performance dashboards displaying rPPG accuracy against local reference standards; and patient-facing messaging that frames rPPG as a screening triage tool rather than a diagnostic replacement. These are governance safeguards proportionate to the safety risk, not optional enhancements.

### 4.5 Limitations for policy interpretation

Distance was self-reported and subject to recall bias; GPS-based distance measures would provide more precise implementation targeting data. Brand was unknown for 21.8% of owners, reducing power for brand contrasts; no brand contrast achieved FDR-adjusted significance, and brand findings should be treated as hypothesis-generating signals. The cross-sectional design precludes causal inference; longitudinal studies are needed to test whether trust calibration interventions alter adoption decisions. Findings are from five sites in Kebbi State, northern Nigeria; device ecology, ownership rates and access burden patterns may differ in other regions, and replication is required before these findings inform national or international policy guidance. Qualitative responses were brief, limiting thematic depth. Future studies should examine whether smartphone ownership, distance from BP services, income, education and connectivity form a broader socioeconomic access gradient that shapes both digital health adoption and perceived usefulness.

## 5 Conclusion

User trust in rPPG screening is structured by digital inclusion and physical access in ways that operate independently of the technology’s validated accuracy. Transsion device familiarity functions as a trust heuristic, while physical access barriers are associated with lower perceived usefulness, together showing that adoption is shaped by device ecology and infrastructure constraints. Health systems considering smartphone-based screening should treat local device ecology, physical access burden, connectivity and trust calibration as core policy and HTA variables—not implementation afterthoughts. In low-resource settings, safe digital health implementation requires not only accurate technology, but also procurement, workflow and governance models aligned with the populations and devices through which care will actually be delivered.

## Data Availability

Anonymised aggregate data supporting the findings are available from the corresponding author on reasonable request. Individual-level data are not publicly archived due to participant privacy constraints.

## Acknowledgements

The authors thank the clinical staff and participants at the study sites in Kebbi State, Nigeria, for their time and cooperation.

## Author contributions

D.D. designed the study, collected data, conducted analyses, and drafted the manuscript.

P.D. supervised the study and critically revised the manuscript. Both authors approved the final version.

## Funding

This research did not receive any specific grant from funding agencies in the public, commercial, or not-for-profit sectors.

## Declaration of competing interests

The authors have no relevant financial or non-financial interests to disclose.

## Supplementary Material

### S1 Supplementary Tables

#### S1.1 Table S1: Firth bias-reduced sensitivity analysis

Firth bias-reduced logistic regression was conducted for brand contrasts among owners to assess sparse-data bias in small brand cells (iPhone *n*=7, Other-Android *n*=10). Results were directionally consistent with the primary HC3-robust models; sign and magnitude were similar throughout; conclusions were unchanged.

**Table S1.**
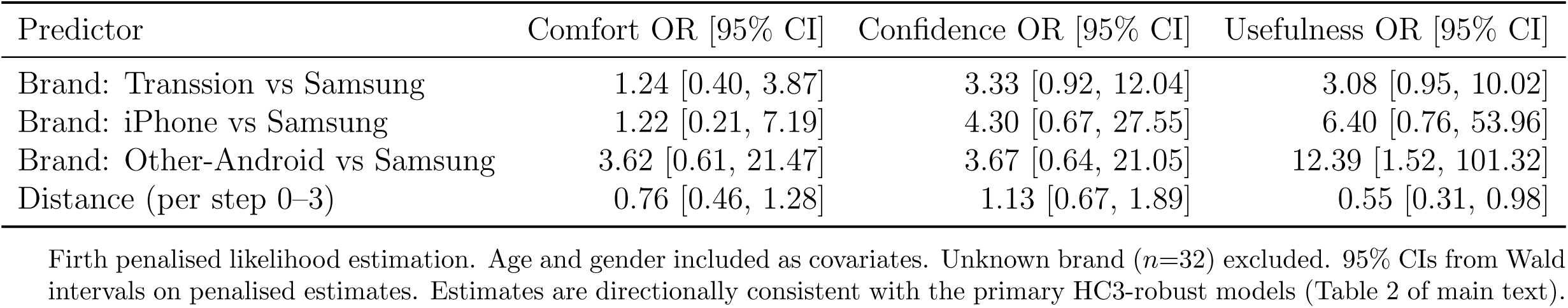
Firth bias-reduced odds ratios for top-box (5/5) outcomes (owners only; Samsung reference)

#### S1.2 Table S2: Benjamini–Hochberg FDR *q* -values for brand contrasts

Main-text *p*-values are descriptive (unadjusted). The table below presents Benjamini–Hochberg (BH) false discovery rate *q*-values computed across the nine brand-contrast tests (three brand comparisons × three outcomes) from the primary top-box logistic models. No brand contrast achieves FDR-adjusted significance (*q <* 0.05), reflecting the limited power of small brand-cell analyses. The unadjusted *p*-values should be interpreted as exploratory implementation signals, not confirmatory evidence of brand-specific effects—a point made explicitly in the main text.

**Table S2.** BH-adjusted *q*-values for brand contrasts (top-box logistic models, owners only; Samsung reference; *N*_tests_=9)

| Outcome | Brand contrast | Unadjusted $p$ | BH $q$ |
| --- | --- | --- | --- |
| Comfort | Transsion vs Samsung | 0.704 | 0.792 |
| Comfort | iPhone vs Samsung | 0.803 | 0.803 |
| Comfort | Other-Android vs Samsung | 0.102 | 0.149 |
| Confidence | Transsion vs Samsung | 0.046 | 0.138 |
| Confidence | iPhone vs Samsung | 0.116 | 0.149 |
| Confidence | Other-Android vs Samsung | 0.116 | 0.149 |
| Usefulness | Transsion vs Samsung | 0.076 | 0.149 |
| Usefulness | iPhone vs Samsung | 0.033 | 0.138 |
| Usefulness | Other-Android vs Samsung | 0.025 | 0.138 |

#### S1.3 Note on ordinal (cumulative logit) sensitivity

Cumulative logit (proportional-odds) models were conducted as a sensitivity analysis for the five-level Likert outcomes. For the brand model (owners only), all three solvers (BFGS, L-BFGS-B, Nelder–Mead) failed to invert the Hessian and did not converge, likely due to near-separation arising from sparse brand cells (iPhone *n*=7, Other-Android *n*=9–10 with Samsung *n*=14 as reference). The top-box logistic models, which are more robust to small cells, are therefore the sole reported results for the brand analysis. Ordinal sensitivity is reported where convergence was achieved; brand contrasts rely entirely on the HC3-robust and Firth logistic estimates (Tables 2 and S1).

### S2 Qualitative Codebook

#### S2.1 Domain definitions

**Digital inclusion:** Android availability, device speed, brand familiarity.

- *Inclusion criteria:* Comments requesting Android versions, faster capture, or referencing specific device brands.
- *Examples:* “make android versions”, “capture should be faster”.
- *Exclusion criteria:* Comments about internet or data costs (coded under Access Constraints); general accuracy comments (Trust Calibration).

**Access constraints:** Travel burden, waiting time, connectivity and data costs, offline-first needs, power reliability.

- *Inclusion criteria:* Comments about internet requirements, data costs, power supply, or travel to a facility.
- *Examples:* “make it work without need for internet”, “Intermittent power supply”.
- *Exclusion criteria:* Device hardware comments (Digital Inclusion); direct accuracy comparisons (Trust Calibration).

**Trust calibration:** Desire for cuff confirmation, perceived accuracy versus convenience, privacy of face video.

- *Inclusion criteria:* Comments about accuracy, validation against standard devices, or privacy concerns about facial capture.
- *Examples:* “improve accuracy above current standard bp device”.
- *Exclusion criteria:* Speed or hardware requests (Digital Inclusion).

Each domain has explicit inclusion and exclusion rules. Two coders independently piloted 20% of entries (*n*≈52), refined the codebook through discussion, then double-coded a random 10% subsample (*n*=26) blinded to quantitative results. The first author coded

the remainder.

### S2.2 Inter-rater reliability (double-coded subsample, *n*=26)

#### S2.3 Core findings by strata

**Ownership:** Owners generated most digital inclusion content (Android availability, device speed, capture affordances), consistent with device access shaping perceived usefulness.

**Table S3.** Inter-rater reliability statistics by domain

S2.2 Inter-rater reliability (double-coded subsample, $n=26$ )
| Domain | $P_o$ | Cohen’s $\kappa$ | Gwet’s AC1 | PABAK |
| --- | --- | --- | --- | --- |
| Digital Inclusion | 0.85 | 0.70 | 0.72 | 0.69 |
| Access Constraints | 0.96 | 0.91 | 0.94 | 0.92 |
| Trust Calibration | 0.73 | 0.46 | 0.47 | 0.46 |

$P_o$ : observed agreement. PABAK: prevalence- and bias-adjusted kappa. Agreement was good–excellent for Digital Inclusion and Access Constraints; moderate for Trust Calibration, reflecting lower base-rate prevalence of trust-calibration codes and hence higher chance-agreement instability.

**Brand group:** Transsion owners concentrated requests on Android build quality, processing speed, flashlight/night-mode capture, and hand placement guidance, aligning with their higher confidence odds in adjusted models. Samsung owners more frequently surfaced accuracy concerns (Trust Calibration). The policy implication is that onboarding and messaging should be tailored to local brand ecology rather than assuming a uniform user experience.

**Distance:** Connectivity and offline-first needs clustered among non-owners and those reporting greater travel distance to a blood pressure facility, consistent with the inverse distance–usefulness association and supporting the policy case for offline-capable workflows.

#### S2.4 Deviant cases

Examples that guard against overly simple narratives: a Samsung owner requested higher accuracy despite acknowledging the convenience of a smartphone approach, while Transsion owners who expressed high enthusiasm still demanded faster capture and better capture affordances—indicating that enthusiasm and critical appraisal coexist and that trust calibration cannot be assumed from ownership or brand alone.

#### S3 Thick Description: Exemplar Quotes

The following verbatim excerpts illustrate the three qualitative domains. Quotes are anonymised; age group and brand ownership are as recorded in the survey.

> *“Make use of device flash light.”*
>
> (Patient, female, 25–34, Transsion owner)
>
> *Domain: Digital Inclusion — platform fit and capture affordances. Relevant to implementation: Transsion-specific flashlight/capture requirements should inform device validation protocols*.
>
> *“Improve accuracy above current standard BP device.”*
>
> (Patient, male, 15–24, Samsung owner)
>
> *Domain: Trust Calibration — explicit accuracy expectation. Relevant to governance: even engaged users set accuracy as a prerequisite for substituting rPPG for cuff measurement*.
>
> *“Make it work without need for internet.”*
>
> (Patient, male, 15–24, brand unknown)
>
> *Domain: Access Constraints — offline-first infrastructure need. Relevant to implementation: offline capability is a recognised requirement, not an optional enhancement*.
>
> *“Make it record BP without stress.”*
>
> (Patient, female, 35–44, brand unknown)
>
> *Domain: Access Constraints — convenience as primary value, consistent with greater travel burden and the desire for an accessible screening alternative*.

These excerpts collectively illustrate that enthusiasm for rPPG is mediated by platform fit (flash/capture affordances), shaped by trust calibration expectations (accuracy versus convenience), and modulated by infrastructure access constraints (offline-first, power reliability).

## Notes

### Competing Interest Statement

The authors have declared no competing interest.

### Author Declarations

Ethical approval was obtained from the Kebbi State Health Research Ethics Committee (107:017/2024) and Bournemouth University Research Ethics Committee (ID 55993).

